# Why is chest CT important for early diagnosis of COVID-19? Prevalence matters

**DOI:** 10.1101/2020.03.30.20047985

**Authors:** Antonio Esposito, Anna Palmisano, Giulia Maria Scotti, Marco J. Morelli, Davide Vignale, Francesco De Cobelli, Giovanni Tonon, Carlo Tacchetti

## Abstract

SARS-CoV-2 viral infection is a global pandemic disease (COVID-19). Reaching a swift, reliable diagnosis of COVID-19 in the emergency departments is imperative to direct patients to proper care and to prevent disease dissemination. COVID-19 diagnosis is based on the identification of viral RNA through RT-PCR from oral-nasopharyngeal swabs, which however presents suboptimal sensitivity and may require several hours in overstressed laboratories. These drawbacks have called for an additional, complementary first line approach. CT is the gold standard method for the detection of interstitial pneumonia, a hallmark feature of COVID-19, often present in the asymptomatic stage of the disease. Here, we show that CT scan presents a sensitivity of 95.48% (std.err=0.35%), vastly outperforming RT-PCR. Additionally, as diagnostic accuracy is influenced by disease prevalence, we argue that predictive values provide a more precise measure of CT reliability in the current pandemics. We generated a model showing that CT scan is endowed with a high negative predictive value (> 90%) and positive predictive value (69 - 84%), for the range of prevalence seen in countries with rampant dissemination. We conclude that CT is an expedite and reliable diagnostic tool to support first line triage of suspect COVID-19 patients in areas where the diffusion of the virus is widespread.

## Introduction

SARS-CoV-2 infection has been recently declared pandemic by the WHO. The associated disease, COVID-19, has been detected in more than 180 countries and territories, with Italy, Iran, USA and Spain experiencing the most widespread outbreaks outside of China.

SARS-CoV-2 infection is highly transmissible, yet COVID-19 has a relatively low death rate (1.0– 3.5%), except in older people (aged >70 years) with comorbidities. It is estimated that 15–20% of infected people develop severe interstitial pneumonia and 5–10% require critical care^1, 2^.

In the absence of specific treatments/vaccines, the most important strategy to save lives is to quarantine people and promptly isolate infected cases. Thus, the ability to obtain an expedite and reliable diagnosis in patients with a clinical suspicion of COVID-19 is imperative, to swiftly identify patients to be directed towards appropriate quarantine or care measures. This is particularly true in the emergency departments (ED), to avoid cross-contamination with non-infected individuals.

Unfortunately, the current protocols often do not warrant a fast and sensitive triage for COVID-19 patients, especially in overstressed regions interested by a steep growth of the epidemic curve. Presently, the diagnosis of COVID-19 is based on RT-PCR identification of SARS-CoV-2 viral RNA (RT-PCR) from oral-nasopharyngeal swab specimens, with lab tests and chest imaging playing ancillary roles. RT-PCR is highly specific (up to 100%) but not equally sensitive (ranging between 30 and 70% depending on viral burden and on the severity of, and time elapsed from symptom onset)^3^. Moreover, the lag time to obtain lab test results may be substantial, depending on various laboratory capabilities^4^.

False-negative results, lag time and yet limited availability of this approach altogether call for an alternative first line diagnostic management, aiming to provide a fast and sensitive diagnosis, while waiting for results provided by RT-PCR screening of nasopharyngeal swabs.

### Computed Tomography (CT) role in early COVID-19 diagnosis: a yin/yang perspective

Computed Tomography (CT) scan represents the gold standard in the diagnosis and characterization of interstitial lung diseases, including viral pneumonia^5^.

Mostly reported COVID-19 CT findings include multifocal ground-glass opacities, affecting both lungs, with peripheral distribution, more frequently involving posterior segments (Figure 1). Bronchovascular thickening is frequently present within the lesion. As the disease progresses, crazy paving with air space consolidation dominates the CT pattern, associated with traction bronchiectasis in more severe patients^6-8^.

**Figure 1.**
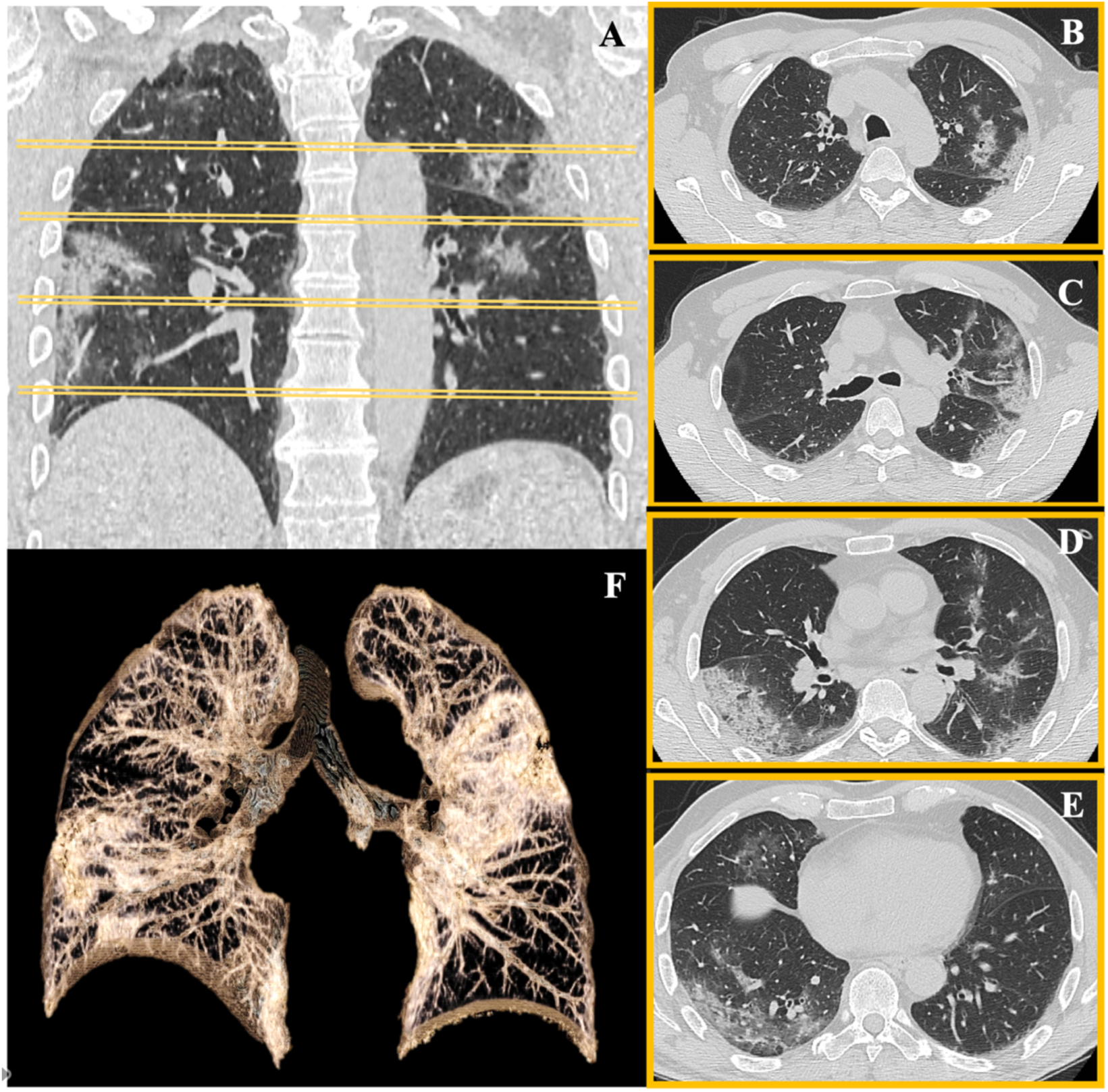
Chest CT in COVID-19 pneumonia. A 61-year-old man from Lodi (endemic city in Lombardy) attended to the emergency department of San Raffaele Hospital in Milan for fever, caught and dyspnoea. Epidemiologic, clinical evaluation and lab tests resulted highly suspicious for SARS-CoV2 infection. Nasopharyngeal swab was collected, and chest CT was immediately performed. CT showed peripheral opacity (A) with ground-glass opacities and crazy-paving pattern involving mainly the upper left lobe (B-D) and the lower right lobe (D-E), with typical posterior involvement. CT findings were highly suggestive for SARS-CoV2 pneumonia. 3D volume rendering CT reconstruction captures the overall pulmonary involvement (F). Results from the first swab (available 24 hours later) resulted negative. Another swab was taken after 3 days, resulting positive.

In our routine daily practice in Lombardy region, the epicenters of COVID-19 epidemics in Italy, CT imaging resulted pervasively altered in COVID-19 patients, even in the early stages of the disease, exactly as widely reported from the recent COVID-19 outbreak in China^6, 9-15^

As a matter of fact, chest CT has been included among the diagnostic criteria during the most rampant phase of epidemic spreading in Wuhan^16^. Notwithstanding, the effectiveness of using CT scan to triage patients with a suspected COVID-19 has been questioned for the matter of specificity, as CT signs of SARS-CoV-2 may partially overlap with other pulmonary viral infections, including influenza, H1N1, SARS and MERS. Hence, the American College of Radiologist (ACR) advised against the use of CT scan as a first line diagnostic tool for patients with suspected COVID-19 infection^17^. Accordingly, the Royal College of Radiologists (RCR) in UK^18^ has advocated against the use of CT scan as a forefront diagnostic tool in this disease.

### A matter of prevalence

In line with our own, and the experience of our Chinese colleagues, we posit that the potential role of CT scan to identify COVID-19 has been undervalued, hampering the identification of false negative RT-PCR cases (Figure 1).

CT outperforms RT-PCR sensitivity in detecting SARS-CoV-2 lung infection^9, 12^. So, why has CT not been included in the first line approach? The main concern is the debated diagnostic specificity. However, most of the currently available literature does not allow to correctly estimate CT specificity, because a negative swab, used as standard of reference, does not exclude the presence of the disease and hence the identification of true negative individuals, as also indicated by the large fraction of the initially RT-PCR-negative (and yet CT-positive) patients actually resulted positive at subsequent swabs^9, 12, 13, 19^.

Moreover, most of the concerns leading to the undervaluation of CT as a first line diagnostic test stem from overrating specificity instead of predictive values. A more informative approach to measure the effective performance of a diagnostic test used to screen a given population, e.g. patients with clinical suspicion of COVID-19, should measure the fraction of true positive and true negative patients correctly identified among all the positive and negative results, hence the Positive Predictive Value (PPV) and the Negative Predictive Value (NPV). PPV and NPV largely depend on the pre-test probability of occurrence for a given disease, which is in turn directly linked to the prevalence of the disease in the given population^20^. In the current scenario, the pre-test probability of having a COVID-19 infection among patients presenting with common symptoms like fever, cough or dyspnea, is exceedingly high in countries facing the epidemic burden.

Therefore, we argue it is crucial to evaluate PPV and NPV for CT scan in the present context of COVID-19 prevalence, in order to assess its potential for the first line diagnostic assessment of suspect COVID-19 patients.

### CT in COVID-19 demonstrates high PPV and NPV values in conditions of high prevalence of the disease

In light of these considerations, we have computed PPV and NPV for CT scan, estimating from literature CT sensitivity and specificity with a meta-analysis (Figure 2) and evaluating different pre-test probabilities (Figure 3).

**Figure 2.**
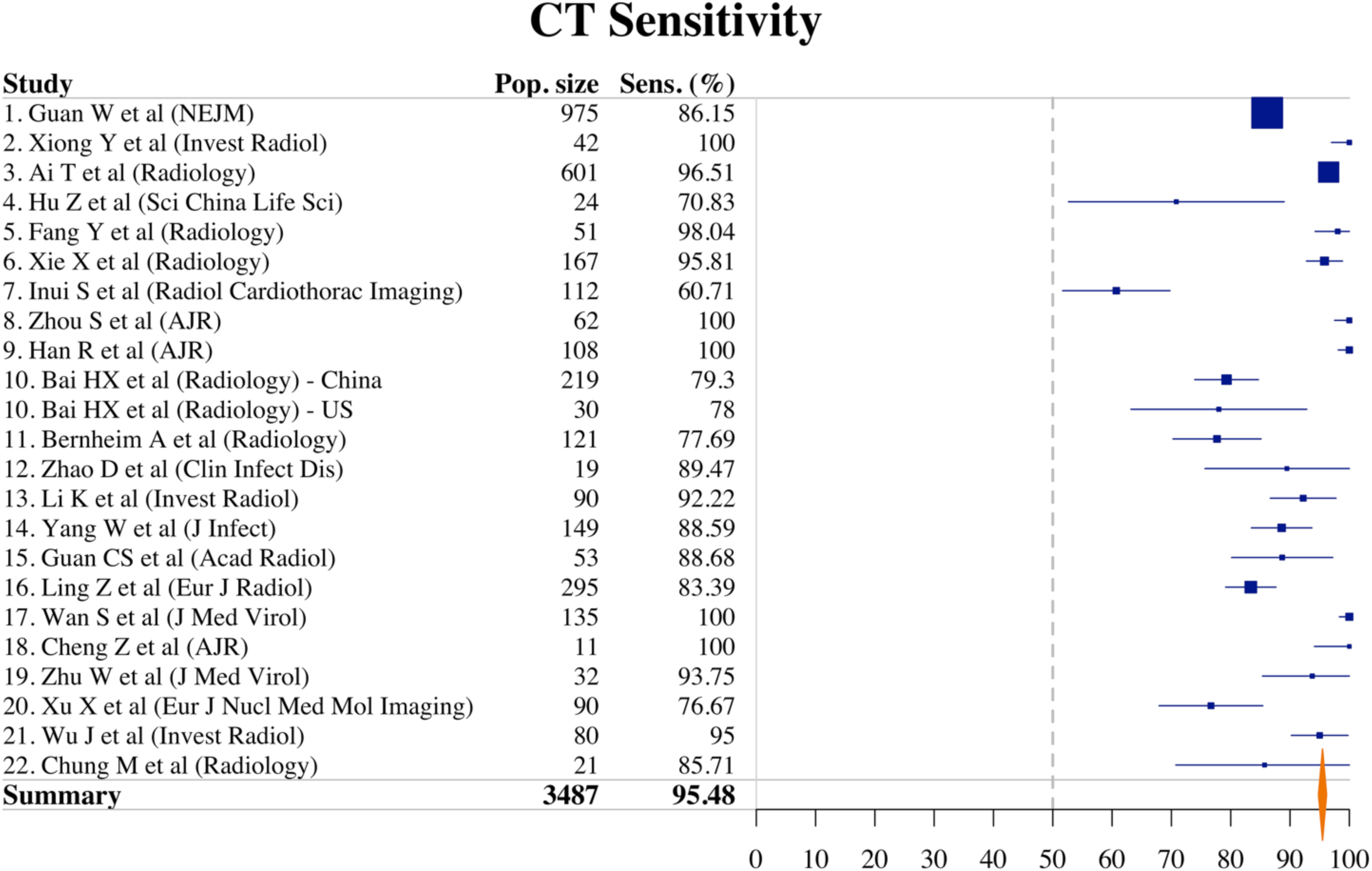
Forest plot from meta-analysis on CT sensitivity. Forest plot from meta-analysis on CT sensitivity in COVID-19 performed on N=22 papers showed a pooled sensitivity of 95.48%. Confidence intervals were estimated with the normal approximation for binomial proportion. For standard error calculations, sensitivity reported in papers 2, 8, 9, 17, 18 was considered as 99%.

**Figure 3.**
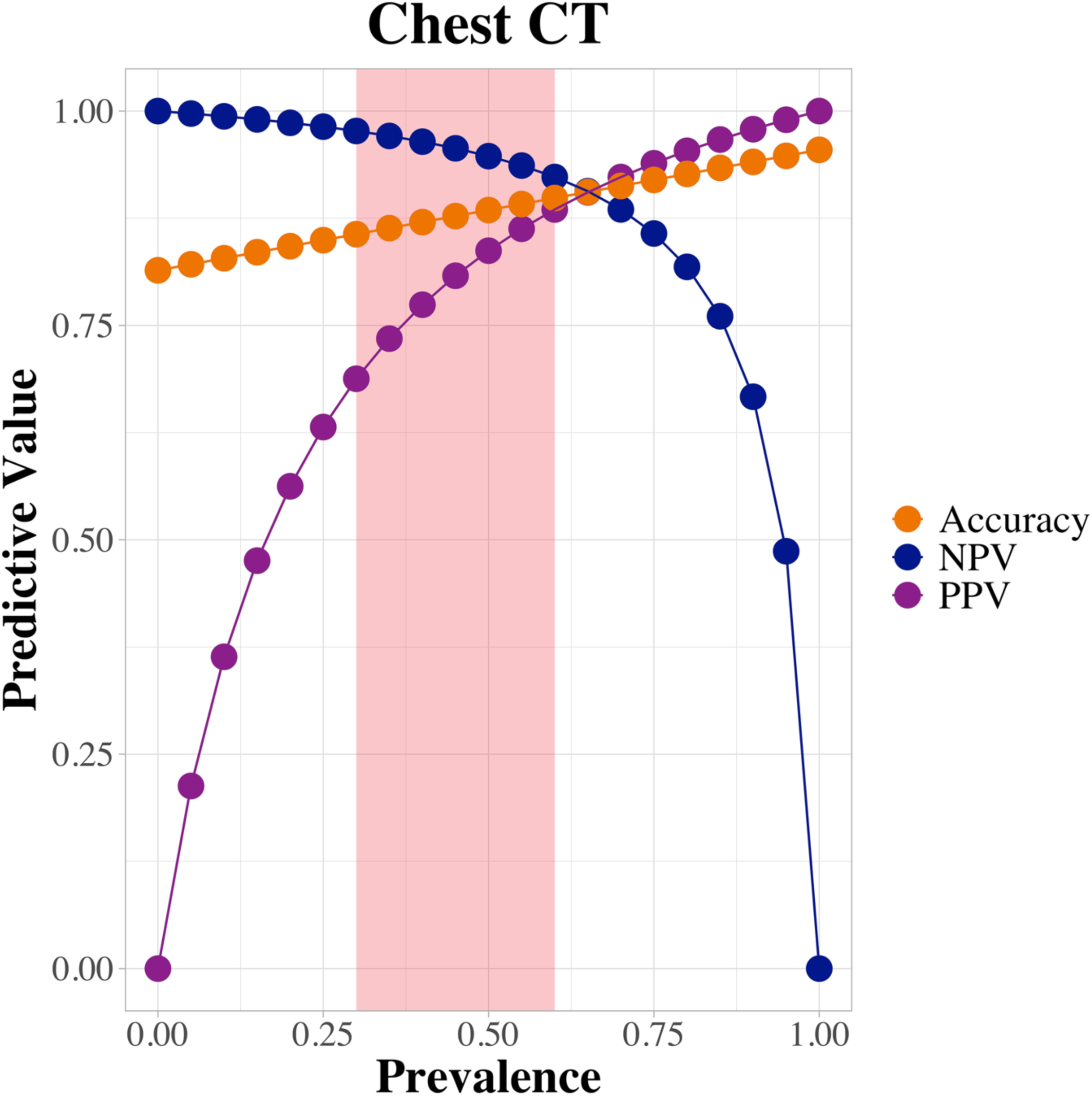
CT predictive values and diagnostic accuracy: prevalence-based modeling. Based on pooled sensitivity and specificity calculated from meta-analysis a model of Negative and Positive Predictive Values (NPV and PPV) and accuracy has been estimated at different prevalence of disease (from 0 to 100%, with 5% interval), with NPV always over 90% between 0% and 65% prevalence of disease. Red column highlights the range of prevalence observed in the areas experiencing rampant phase of epidemic spreading.

More in detail, N=120 reports were identified through PUBMED database searching and screened. The included publications needed to be in English language and include COVID-19 patients diagnosed with RT-PCR and reporting CT findings and performances. Case reports, commentary and review were excluded, as well as studies on pediatric populations and pregnant women, leaving N=22 studies^9-^15, 21^-35^ (Table 1). The included studies had fair and good quality scores according to “The NIH Quality Assessment Tool for Case Series Studies”^36^, sharing clear study design and aims, comparable population, clear measurement of outcome and clear description and/or representation of results. Since most of the selected studies were conducted on COVID-19 RT-PCR-confirmed positive patients, estimates about CT specificity were challenging to retrieve. Among the four studies comparing CT in COVID-19 and non-COVID-19 pneumonia^23, 25, 31, 32^, only data from Bai HX et al.^23^ provide a reliable estimation of CT specificity, since it is the only study assessing performance in distinguishing COVID-19 (219 patients) from other ascertained viral interstitial pneumonia (205 patients). Differently, in the remaining studies only negativity to swab is taken into account to target pneumonia as non-COVID-19, and often the same authors claim doubts about the actual final diagnosis. Hence, considering that Bai HX et al.^23^ reported the performance of 7 independent radiologists, we estimated CT specificity=81.43% (std.err=2.38%) as the pooled CT specificity of the three Chinese and the four American radiologists.

**Table 1.** Main characteristics of studies evaluating diagnostic performances of CT in COVID-19 pneumonia.

| Study | Quality | Study design | Cohort size (N) | Patients age (Y.O.) | RT-PCR Indicative for COVID-19 (N) | Chest CT suggestive for COVID-19 (N) | Sensitivity* %, N/D (95% CI) | RT-PCR Indicative for NON COVID-19 virus (N) | Chest CT suggestive for NON COVID-19 (N) | Chest CT negative for pneumonia | Specificity* %, N/D (95% CI) | Number of CT readers |
| --- | --- | --- | --- | --- | --- | --- | --- | --- | --- | --- | --- | --- |
| **1.**<br>Guan W et al (NEJM) | Good | Retrospective observational Cohort | 1099 | 47° | 1099 (975 with CT scan at the time of admission) | 840 | 86.15%, 840/975 (83.98%-88.32%)† | NA | NA | 135 | NA | 2/3 |
| **2.**<br>Xiong Y et al (Invest Radiol) | Good | Retrospective observational Cohort | 42 | 26-75^ | 42 | 42 | 100%, 42/42 (96.99%-100.00%)† | NA | NA | 0 | NA | 3 |
| **3.**<br>Ai T et al (Radiology) | Good | Retrospective observational Cohort | 1014 | 51<sup>§</sup> | 601 | 580 | 96.51% 580/601 (95.04%-97.98%)† | CD | NA | 126 | NA | 2 |
| **4.**<br>Hu Z et al (Sci China Life Sci) | Fair | Retrospective observational Cohort | 24 | 32.5° | 24 | 17 | 70.83%, 17/24 (52.64%-89.02%)† | NA | NA | 7 | NA | NR |
| 5.<br>Fang Y et al<br>(Radiology) | Fair | Retrospective<br>observational<br>Cohort | 51 | 45° | 51 | 50 | 98.04%%,<br>50/51<br>(94.24%-100%)† | NA | NA | 1 | NA | NR |
| 6.<br>Xie X et al<br>(Radiology) | Fair | Retrospective<br>observational<br>Cohort | 167 | NR | 167 | 160 | 95.81%, 160/167<br>(92.77%-<br>98.85%)† | NA | NA | 7 | NA | 2 |
| 7.<br>Inui S et al<br>(Radiol<br>Cardiothorac<br>Imaging) | Good | Retrospective<br>observational<br>Cohort | 112 | 25-93^ | 112 | 68 | 60.71%,<br>68/112<br>(51.66%-<br>69.76%)† | NA | NA | 44 | NA | 3 |
| 8.<br>Zhou S et al<br>(AJR) | Good | Retrospective<br>observational<br>Cohort | 62 | 30-77^ | 62 | 62 | 100%,<br>62/62<br>(97.52%-100%)† | NA | NA | 0 | NA | 2 |
| 9.<br>Han R et al<br>(AJR) | Good | Retrospective<br>observational<br>Cohort | 108 | 21-90^ | 108 | 108 | 100%,<br>108/108<br>(98.12%-100%)† | NA | NA | 0 | NA | 2 |
| 10.<br>Bai HX et al<br>(Radiology) | Good | Retrospective<br>observational<br>Cohort | 424 | 3-96^ | 219 | 158, 157,<br>206 for each<br>Chinese<br>radiologist | 72.15%, 71.69%,<br>94.06% for each<br>radiologist,<br>average=79.3%<br>(73.93%-<br>84.67%)† | 205 | 192, 181, 49<br>for each<br>Chinese<br>radiologist | 0 | 93.66%, 88.29%,<br>23.9% for each<br>radiologist,<br>average=68.62%<br>(62.27%-74.97%)† | 3 |
|  |  |  | 58 |  | 30 | 28, 25, 22,<br>21 for each<br>American<br>radiologist | 93.33%, 83.33%,<br>73.33%, 70% for<br>each radiologist,<br>average=78%<br>(63.18%-<br>92.82%)† | 28 | 28, 26, 26,<br>28 for each<br>American<br>radiologist | 0 | 100%, 92.86%,<br>92.86%, 100% for<br>each radiologist,<br>average=96.43%<br>(89.56%-100%)† | 3+4 |
| 11. Bernheim<br>A et al<br>(Radiology) | Good | Retrospective<br>observational<br>Cohort | 121 | 18-80^ | 121 | 94 | 77.69%,<br>94/121<br>(70.27%-<br>85.11%)† | NA | NA | 27 | NA | 2 |
| **12.**<br>Zhao D et al<br>(Clin Infect Dis) | Fair | Retrospective observational Cohort | 34 | 48 / 35<br>(median in COVID / non-COVID patients) | 19 | 17 | 89.47%,<br>17/19<br>(75.67%-100%)† | CD | NA | 0 | NA | NR |
| **13.**<br>Li K et al<br>(Invest Radiol) | Fair | Retrospective observational cohort | 90 | 45.5<sup>§</sup> | 90 | 83 | 92.22%,<br>83/90<br>(86.69%-97.75%)† | NA | NA | 7 | NA | 2 |
| **14.**<br>Yang W et al (J Infect) | Good | Retrospective observational Cohort | 149 | 45.1<sup>§</sup> | 149 | 132 | 88.59%,<br>132/149<br>(83.49%-93.69%)† | NA | NA | 17 | NA | 2 |
| **15.**<br>Guan CS et al<br>(Acad Radiol) | Good | Retrospective observational Cohort | 53 | 42<sup>§</sup> | 53 | 47 | 88.68%,<br>47/53<br>(80.15%-97.21%)† | NA | NA | 6 | NA | 2 |
| **16.**<br>Ling Z et al<br>(EJR) | Fair | Retrospective observational Cohort | 295 | NR | 295 | 246 | 83.39%,<br>246/295<br>(79.14%-87.64%)† | NA | NA | 49 | NA | NR |
| **17.**<br>Wan S et al (J Med Virol) | Fair | Observational Cohort | 135 | 47 ° | 135 | 135 | 100%,<br>135/135<br>(98.32%-100.00%)† | NA | NA | 0 | NA | NR |
| **18.** Cheng Z et al (AJR) | Good | Retrospective observational Cohort | 33 | 45.8<sup>§</sup> | 11 | 11 | 100%,<br>11/11<br>(94.12%-100.00%)† | CD | NA | 2 | NA | 3 |
| **19.**<br>Zhu W et al (J Med Virol) | Good | Retrospective observational Cohort | 116 | 27-53<sup>^</sup> | 32 | 30 | 93.75%,<br>30/32<br>(85.36%-100.00%)<sup>†</sup> | CD | NA | 30 | NA | 2 |
| **20.**<br>Xu X et al (Eur J Nucl Med Mol Imaging) | Good | Retrospective observational Cohort | 90 | 18-86<sup>^</sup> | 90 | 69 | 76.67%,<br>69/90<br>(67.93%-85.41%)<sup>†</sup> | NA | NA | 21 | NA | 2 |
| **21.**<br>Wu J et al (Invest Radiol) | Good | Retrospective observational Cohort | 80 | 15-79<sup>^</sup> | 80 | 76 | 95%,<br>76/80<br>(90.22%-99.78%)<sup>†</sup> | NA | NA | 4 | NA | 2 |
| **22.** Chung M et al (Radiology) | Good | Retrospective observational Cohort | 21 | 29-77<sup>^</sup> | 21 | 18 | 85.71%,<br>18/21<br>(70.74%-100.00%)<sup>†</sup> | NA | NA | 3 | NA | 2 |

NR = not reported, CD = cannot determine, NA = not applicable.
<sup>°</sup>Data are median value.
<sup>§</sup>Data are mean value.
<sup>^</sup>Data are range.
\* Data are percentage, with numerators and denominators in parenthesis. Data in brackets are 95% Confidence Intervals (CIs).
<sup>†</sup> Estimated values. CIs were estimated with the Normal approximation for binomial proportion.

Similarly, for CT sensitivity, we performed a meta-analysis applying the generic inverse variance method^37^ as implemented in the *meta* Rpackage, with standard errors inferred by normal approximation of the binomial distribution, obtaining the following pooled estimates: sensitivity=95.48% (std.err=0.35%) (Figure 2).

These data were used to evaluate the CT positive (PPV) and negative (NPV) predictive values and CT accuracy at different prevalence of disease (from 0 to 100% prevalence, with 5% interval) (Figure 3).

As expected, all the three measures of CT scan preformance were significantly influenced by the prevalence of the disease (Figure 3). Notably, CT scan guarantees a very high negative predictive value (NPV), higher than 90%, for prevalence up to 65%. This is keenly relevant especially in regions with rapidly escalating spread of the infection. According to the public data provided by the Italian Civil Protection Agency^38^, we have estimated a 39% prevalence of COVID-19, as of March 29th 2020, among suspected patients (ratio between affected patients on number of RT-PCR) in the Lombardy region. This proportion is very similar to the fraction of affected/screened patients reported in Wuhan, China^9, 39^ and represents a reliable estimate of the pre-test probability of patients submitted to screening. Notably, with a pre-test probability ranging from 30 to 50%, the interval currently seen in various countries facing COVID-19 epidemics, the NPV of CT scan was between 97.68% and 94.74%, the PPV between 68.78% and 83.72%, and the accuracy between 85.65% and 88.46% respectively (Figure 3). These data hence suggest that CT represents a valuable tool for the accurate initial triage of patients in the congested hospitals facing the COVID-19 epidemics, with a remarkably high NPV and an acceptable PPV. The few expected false positive cases at CT scan represent a minor problem if compared to the risk of missing or delaying the diagnosis of COVID-19, and include patients affected by other forms of interstitial lung diseases rather than healthy individuals.

Importantly, CT scan may identify lung abnormalities from COVID-19 infection in a relevant amount (54%) of asymptomatic patients, according to a recent study on cases from the cruise ship Diamond Princess^22^, as well as among patients with symptoms highly suggestive of COVID-19 and with a yet negative RT-PCR^9, 12, 13, 19^.

### CT is a reliable, swift diagnostic tool for the early identification of COVID-19 infection in endemic areas

All together, these data indicate high diagnostic accuracy of CT at the prevalence rate currently seen in most countries where the COVID-19 epidemic is rising or established. As such, CT scan may identify patients affected by COVID-19 in the critical stages of epidemic spreading, when it is crucial to reach prompt clinical decisions and where oral-nasopharyngeal swab specimens may not provide yet trustworthy and rapid information. CT scan can be performed and interpreted in a handful of minutes^40^. Moreover, the recent introduction of artificial intelligence (AI) approaches as diagnostic tools for semi-automated COVID-19 CT scan reporting may even shorten this time and improve performances^41^.

Thus, CT integration in the diagnostic process may help for an expedite and reliable diagnosis of COVID-19 in areas where the diffusion of the virus is rampant. As CT scan resources are widely available in many countries, we suggest that this approach should be taken into account in the planning of health care procedures to withstand the current pandemic expansion.

## Data Availability

All data are included in the embedded Table 1.

